# GFAP as a Biomarker of Stroke Subtype and Brain Tissue Damage in Acute Stroke: A Systematic Review and Meta-Analysis

**DOI:** 10.64898/2026.09.11.26362885

**Authors:** Catherine Brassard, Julien F. Paul, Daniela Ziegler, Miguel Chagnon, Olena Bereznyakova, Mark Keezer, Laura C. Gioia

**Affiliations:** Department of Neurosciences, Université de Montréal; Faculty of Medicine, Université de Montréal; Division of Neurology, CHUM; Direction de l’Enseignement et de l’Académie, CHUM; Department of Mathematics and Statistics, Université de Montréal; Neurosciences Axis, Centre de Recherche du CHUM (CRCHUM)

**Keywords:** Acute Stroke, Ischemic Stroke, Intracerebral Hemorrhage, Blood Biomarkers, Glial Fibrillary Acidic Protein

## Abstract

**Background:** Glial fibrillary acidic protein (GFAP) is a promising blood biomarker that can distinguish intracerebral hemorrhage (ICH) from acute ischemic stroke (AIS). We aimed to summarize the existing evidence regarding: 1) the diagnostic accuracy of GFAP to differentiate ICH from AIS, and from undifferentiated suspected stroke, and 2) characterize GFAP’s diagnostic accuracy across time strata from stroke onset. As a secondary objective, associations between GFAP and clinical and radiological outcomes were narratively summarized.

**Methods:** Five databases [(MEDLINE/EMBASE/EBM Reviews/CINAHL Complete/Google Scholar] were searched (03/2026) for studies reporting blood GFAP levels in acute stroke. GFAP levels were analyzed according to time from onset to sampling. Risk of bias was assessed using QUADAS-2 for studies reporting diagnostic accuracy data. Forest plots, pooled sensitivities and specificities were calculated.

**Results:** Two reviewers independently screened 6,336 studies; 64 were included for systematic review and 18 for meta-analysis. Risk of bias was low overall among diagnostic studies. For differentiating ICH from AIS, pooled AUC was 0.86 (95% CI: 0.76-0.92), pooled sensitivity 74.4% (95% CI:63.2-84.2%) and pooled specificity 87.2% (95% CI:77.0-94.7%). For ICH versus undifferentiated suspected stroke, pooled AUC was 0.88 (95% CI: 0.79-0.95), pooled sensitivity 75.4% (95% CI: 63.6–83.3%) and pooled specificity 90.5% (95% CI:88.0-96.2%). Diagnostic precision improved in earlier time strata: <6h: AUC 0.91, sensitivity 75.0%, specificity 92.7%; <2h: AUC 0.96, sensitivity 76.7%, specificity 96.4%. GFAP correlated with ICH volume in 11/12 studies, although smaller hematomas (<10 mL) were frequently associated with GFAP values below limits of detection. Few studies (n=3) suggest higher GFAP levels correlate with greater ischemic injury in AIS.

**Conclusions:** As a blood biomarker in acute stroke, GFAP has strong rule-in value (i.e. high specificity) for ICH, which is preserved in earlier time windows. Nevertheless, sensitivity remains limited, restricting its use as a rule-out test, particularly for smaller ICH volumes. Knowledge gaps remain regarding GFAP’s potential in assessing ischemic injury extent in AIS.

## INTRODUCTION

Acute stroke is classified according to the underlying pathophysiology. Ischemic stroke, caused by a vessel occlusion, accounts for the majority (70-85%) of stroke cases, while intracerebral hemorrhage (ICH), due to vessel rupture, represents 15-20% of cases with regional variation.(1) The concept “*time is brain*” emphasizes that every minute of treatment delay results in an estimated loss of 1.9 million neurons in severe stroke.(2) Rapid identification of stroke subtype is necessary to administer timely disease-specific treatments, such as thrombolysis and/or thrombectomy in acute ischemic stroke (AIS) and early initiation of blood pressure and anticoagulant reversal to limits hematoma growth in ICH.(3,4) Diagnosing acute stroke and its subtype requires brain neuroimaging (computed tomography (CT) and/or magnetic resonance imaging (MRI)).(5) In the case of AIS of delayed presentation or unknown onset, advanced neuroimaging such as CT perfusion is also necessary to determine eligibility for acute treatments.(5–7)

Brain-specific biomarkers are promising adjunct tools that could expedite diagnosis of acute stroke subtype, particularly in out-of-hospital settings where neuroimaging is not readily available. Glial fibrillary acidic protein (GFAP), a highly brain-specific astrocytic intermediate filament protein, has been studied extensively for this purpose.(8) In the acute phase of stroke, circulating GFAP concentrations are significantly higher in ICH compared to AIS,(9) presumably due to a rapid increase in blood GFAP levels following abrupt disruption of the blood-brain-barrier caused by hematoma formation and direct destruction of the surrounding astrocytes. In contrast, GFAP levels appear to rise more gradually in AIS, presumably due to the delayed astroglial cell death as ischemic penumbral tissue evolves into infarct core over time in the absence of reperfusion.(10)

Prior studies indicate that GFAP demonstrates strong diagnostic performance for differentiating ICH from AIS within 6 hours from stroke onset.(9,11,12) Beyond stroke subtype differentiation, GFAP concentrations have also been associated with infarct volumes (13–15), supporting its potential utility in AIS of unknown onset.(16) Point-of-care devices capable of measuring and quantifying GFAP concentrations within minutes are in development, bringing this biomarker closer to real-time clinical application.(17) Ultimately, if stroke subtype (AIS vs. ICH) and the extent of brain tissue injury can be accurately estimated using point-of-care tools, disease-specific management could be expedited and extended to prehospital and resource-limited settings.(11,18,19) This systematic review and meta-analysis has 3 objectives: 1) provide an updated synthesis of the diagnostic accuracy of blood GFAP in acute stroke for differentiating ICH from AIS and from undifferentiated suspected stroke, and 2) characterize the available evidence regarding diagnostic accuracy of GFAP across time strata from stroke onset. As a secondary, objective, we narratively summarized the associations between GFAP concentrations and clinical outcomes (stroke severity as defined by the NIHSS), and radiological outcomes (ASPECTS scores, ICH volumes).

## METHODS

### Study search and selection

This systematic review was conducted in accordance with the Preferred Reporting Items for Systematic Reviews and Meta-Analyses for Diagnostic Test Accuracy Studies (PRISMA-DTA) guidelines, with the protocol registered on PROSPERO (CRD42024608605).(20) In collaboration with a health sciences librarian (DZ), four electronic bibliographic databases [MEDLINE (OVID), Embase (OVID), EBM Reviews (OVID), and CINAHL Complete (EBSCO)] were systematically searched, along with Google Scholar and 3 grey literature sources (ClinicalTrials.gov, the World Health Organization International Clinical Trials Registry Platform [ICTRP], and the International Standard Randomised Controlled Trial Number [ISRCTN] registry, from inception to August 2024. An updated search through March 2026 was conducted to incorporate relevant studies published after the initial search. No date restrictions were applied. Search strategies were adapted for each database and included the following key words and their combinations: “glial fibrillary acidic protein”, “GFAP”, “stroke”, “cerebrovascular accident”, “intracerebral hemorrhage”, “ischemic stroke”, and “transient ischemic attack’’. The full search strategy is published online: https://doi.org/10.5683/SP3/WBDI4L.

Two authors (CB, JFP) independently screened titles and abstracts of all identified records. From these, full texts of potentially eligible articles were subsequently retrieved and assessed for study inclusion. Eligible studies included original studies evaluating blood GFAP sampling in participants with acute suspected stroke after excluding duplicates using EndNote 21 and Covidence tools. Discrepancies between reviewers were resolved by consensus, and, where necessary, by consultation with a third reviewer (LCG). Authors of relevant studies were contacted when full text and/or necessary information for the systematic review were required for the meta-analysis.

### Selection criteria

Included studies reported recruitment of adult participants (≥18 years) presenting with a suspected acute stroke and diagnosed as either AIS, ICH, transient ischemic attack (TIA) or stroke mimic according to reference standards, in whom blood GFAP levels were measured, regardless of time of onset (known or unknown). Studies were excluded if: 1) the primary focus of the study included animal models or cell cultures, 2) participants were <18 years of age, 3) circulating GFAP levels were measured elsewhere than in blood (e.g. cerebrospinal fluid, urine), 4) they consisted of abstracts, case-reports, reviews, letters or editorials. For the meta-analysis, studies evaluating the diagnostic accuracy of GFAP were included if sensitivity, specificity and the number of events were reported.

### Data extraction

CB and JFP independently extracted data using Covidence and customized templates in REDCap, a secure web-based application. The following information was extracted: study characteristics (author, year of publication, title, and study design), details of study population (sample size, age, sex, history of stroke), and diagnosis information, and type of neuroimaging performed, stroke subtype, time of stroke onset or last known well [LKW]), median National Institutes of Health Stroke Scale (NIHSS) score, median Alberta Stroke Program Early CT Score (ASPECTS) score, and median modified Rankin scale (mRS) score). Information regarding GFAP measurements (assay method, units, time from stroke onset to sampling or time from LKW to sampling) as well as diagnostic accuracy metrics (cut-off values, true positives [TP], false positives [FP], true negatives [TN], false negatives [FP], area under the curve [AUC], sensitivity, specificity) were also extracted. When necessary, GFAP values were converted to ng/mL for uniformity across studies. Discrepancies in data extraction were resolved using the same consensus process described above. A meta-analysis was performed to assess the diagnostic accuracy of GFAP to distinguish ICH from AIS and from undifferentiated suspected stroke. Data regarding GFAP sensitivity and specificity, TP, TN, FP, FN, and time from onset to blood sampling were used to conduct analyses regarding GFAP’s diagnostic performance.

### Quality assessment

Risk of bias was assessed independently by the two authors (CB, JFP) using the quality assessment tool (QUADAS-2) for studies reporting diagnostic accuracy data (i.e. sensitivity, specificity, or area under the curve). Four domains were evaluated for each study: participant selection, index test(s), reference standard, as well as flow and timing. In case of disagreement, studies were reviewed and discussed with a third author (LCG) to establish consensus. Findings from studies contributing only descriptive or associative data such as correlations with clinical (NIHSS) or radiological outcomes (ASPECTS scores, ICH volumes) were narratively summarized.

### Statistical analysis

Statistical analyses were performed with RStudio (version 2024.04.2+764) and IBM SPSS Statistics (version 29.0.2.0). Data are expressed as absolute percentage (%) for categorical variables, mean ± standard deviation (SD) for normally distributed variables and median and interquartile range (IQR) for non-normally distributed variables. Where necessary, TP, TN, FP and FN counts were back-calculated from the extracted sensitivity and specificity together with the number of individuals with and without ICH when raw 2×2 counts were not directly reported. Derived counts were rounded to the nearest integer. Confidence intervals for GFAP sensitivity and specificity were then calculated from these TP, TN, FP and FN using the Clopper-Pearson exact method. GFAP pooled sensitivity, pooled specificity and area under the hierarchical summary receiver operating characteristic (HSROC) curve were calculated using the *meta (8.5.0)* and *mada (0.5.12)* packages in R, accounting for between-study variation in thresholds by modeling correlations between sensitivity and specificity. A random-effects model was applied, and an arcsine-based transformation enabled stabilization of studies’ variances. Between-study heterogeneity was assessed using the Zhou-Dendukuri bivariate I². Publication bias was evaluated using Deeks’ funnel plot asymmetry test where feasible (≥10 studies, p<0.10 considered significant).

## RESULTS

### 1. Search results

A total of 6,336 studies were identified with the search strategy, of which 6,178 were screened after removal of duplicates. Of these, 64 studies met inclusion criteria for the systematic review. (Figure 1) Eighteen studies reported diagnostic accuracy data sufficient for pooling and were included in the meta-analysis.(Table 1) Seventeen studies only reported associations between GFAP concentrations and clinical (NIHSS) and/or radiological outcomes (ASPECTS, ICH volume). The remaining 29 studies did not report diagnostic accuracy data or outcome association data eligible for synthesis; specific reasons for each study are reported in Supplementary Table 1.

**Figure 1:**
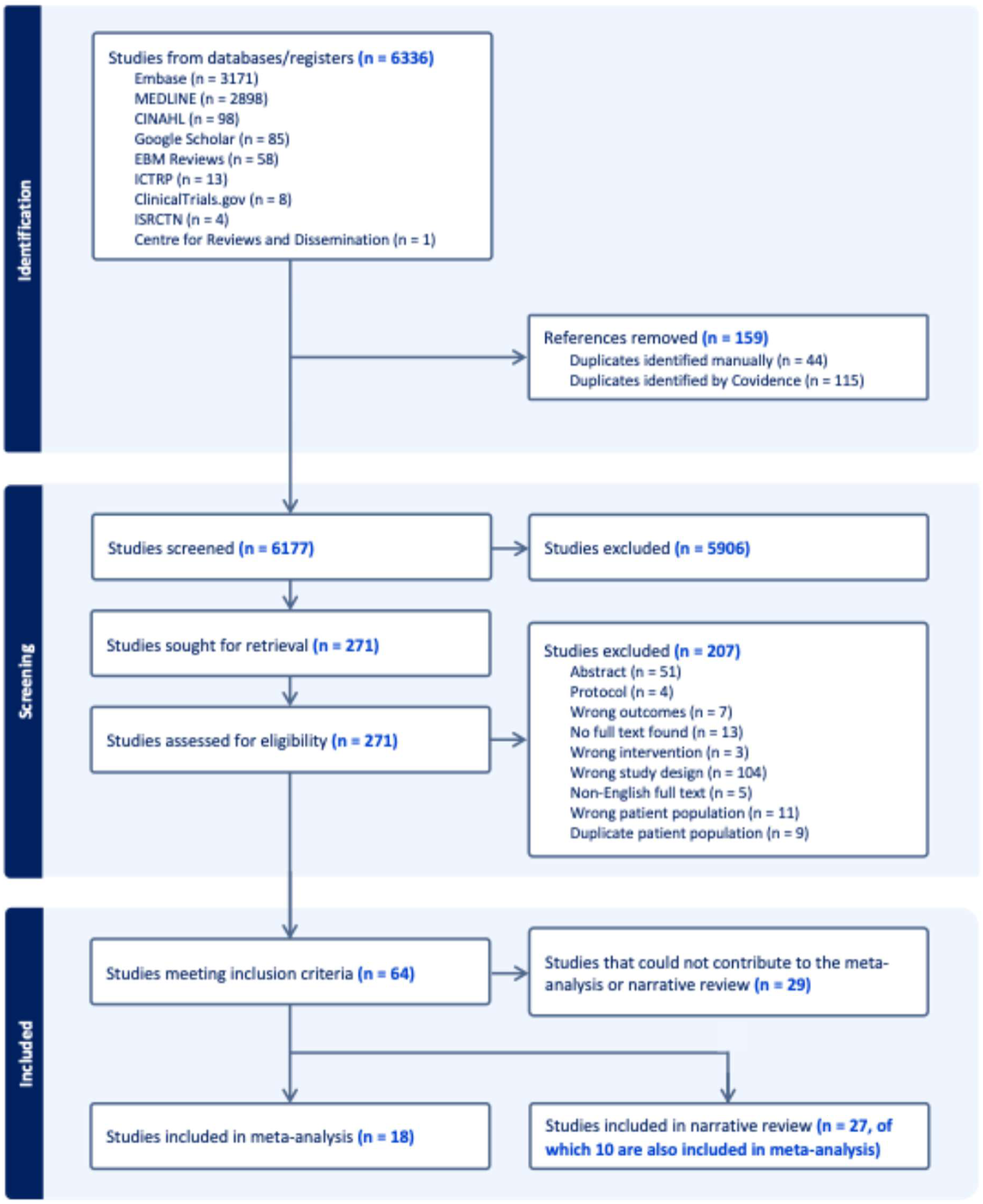
PRISMA-DTA Flow Diagram.

**Table 1:** Characteristics of the 18 studies included in the meta-analysis.

| Author | Year | Study design | Diagnostic Comparison | Study population | Time from onset to sampling | GFAP measurement method | Reference standard | Cut-off (ng/mL) | Cut-off selection |
| --- | --- | --- | --- | --- | --- | --- | --- | --- | --- |
| Bhatia | 2020 | Prospective cohort study | ICH vs AIS | AIS = 187<br>ICH = 63 | <24 h | ELISA | CT, MRI | 0.300 |  |
| Foerch | 2006 | Prospective cohort study | ICH vs AIS | AIS = 93<br>ICH = 42 | <6 h | ECL | CT, MRI | 0.003 | ROC analysis from individual study data |
| Foerch | 2012 | Prospective cohort study | ICH vs. undifferentiated suspected stroke | AIS = 163<br>ICH = 39<br>SM = 3 | <4.5 h | SIMOA | CT, MRI | 0.290 | According to previous study |
| Han | 2023 | Retrospective cohort study | ICH vs. AIS | AIS = 51<br>ICH = 22 | <6 h | SIMOA | CT, MRI | 0.703 | ROC analysis from individual study data |
| Jeager | 2023 | Prospective cohort study | ICH vs undifferentiated suspected stroke | AIS = 131<br>ICH = 21<br>SM = 114<br>TIA = 22 | <4.5 h | SIMOA | CT, MRI | 0.838 | ROC analysis from individual study data |
| Kalra | 2021 | Prospective cohort study | ICH vs undifferentiated suspected stroke | AIS = 75<br>ICH = 70<br>SM = 10 | <12 h | SIMOA | CT | 0.570 | ROC analysis from individual study data |
| Kalra | 2025 | Prospective cohort study | ICH vs undifferentiated suspected stroke | AIS = 258<br>ICH = 76<br>SM = 19 | <6 h | ELISA | CT | 0.055 | ROC analysis from individual study data |
| Katsanos | 2017 | Prospective cohort study | ICH vs AIS | AIS = 121<br>ICH = 34<br>SM = 31<br>Controls = 79 | <6 h | BAT |  | 0.430 | ROC analysis from individual study data |
| Llombart | 2016 | Retrospective cohort study | ICH vs. AIS | AIS = 38<br>ICH = 28 | <6 h | ELISA | CT | 0.070 | ROC analysis from individual study data |
| Author (Cont) | Year | Study design | Diagnostic Comparison | Study population | Time from onset to sampling | GFAP measurement method | Reference standard | Cut-off (ng/mL) | Cut-off selection |
| Luger | 2017 | Prospective cohort study | ICH vs undifferentiated suspected stroke | AIS = 146<br>ICH = 45<br>SM = 11 | <6 h | ELISA | CT, MRI | 0.030 | ROC analysis from individual study data |
| Luger | 2020 | Prospective cohort study | ICH vs undifferentiated suspected stroke | AIS = 171<br>ICH = 64<br>SM = 16 | <6 h | ECL | CT, MRI | 0.072 | ROC analysis from individual study data |
| Nunez-Jurado | 2025 | Prospective cohort study | ICH vs. AIS | AIS = 344<br>ICH = 44 | <6 h | Proteomic assays | CT, MRI |  |  |
| Popa | 2025 | Prospective cohort study | ICH vs AIS | AIS = 37<br>ICH = 32<br>TIA = 16 | <6 h | CMIA | CT | 0.077 | ROC analysis from individual study data |
| Ren | 2016 | Prospective cohort study | ICH vs. AIS | AIS = 79<br>ICH = 45<br>TIA = 3<br>Controls = 57 | <24 h | ELISA | CT, MRI | 0.340 | ROC analysis from individual study data |
| Rozanski | 2016 | Prospective cohort study | ICH vs. AIS | AIS = 49<br>ICH = 25 | Median: 1.04<br>(0.60-2.32) | ECL | CT | 0.290 | According to previous study |
| Stanca | 2015 | Prospective cohort study | ICH vs AIS | AIS = 49<br>ICH = 23<br>Controls = 52 | <12 h | ELISA | CT |  |  |
| Uden | 2009 | Prospective cohort study | ICH vs. AIS | AIS = 83<br>ICH = 14 | <24 h | ELISA | CT, MRI | 0.040 | ROC analysis from individual study data |
| Xiong | 2015 | Prospective cohort study | ICH vs. AIS | AIS = 65<br>ICH = 43 | Range: 2-6 h | ELISA | CT, MRI | 0.700 | ROC analysis from individual study data |

Legend: AIS = Acute ischemic stroke; BAT = Biochip Array Technology; CMIA = Chemiluminescent immunoassay; CT = Computed tomography; ECL = Electrochemiluminescence; ELISA = Enzyme-Linked Immunosorbent Assay; ICH = Intracerebral hemorrhage; MRI = Magnetic resonance imaging; ROC = Receiver-operating characteristic; SIMOA = Single Molecule Array; SM = Stroke Mimics; TIA = Transient Ischemic Attack

### 2. Baseline study characteristics

#### a. Risk of Bias

Across the 18 studies reporting diagnostic accuracy data and included in the meta-analysis, 13 had a low risk of bias across all 4 domains, whereas 3 had an unclear risk for patient selection, (20–22) one for reference standard (23) and another for flow and timing.(24) (Figure 2). Four studies had unclear concerns regarding applicability, involving patient selection in 3 studies (20,22,25) and the index test in one (26), while another study had high applicability concerns related to the index test (27).

**Figure 2:**
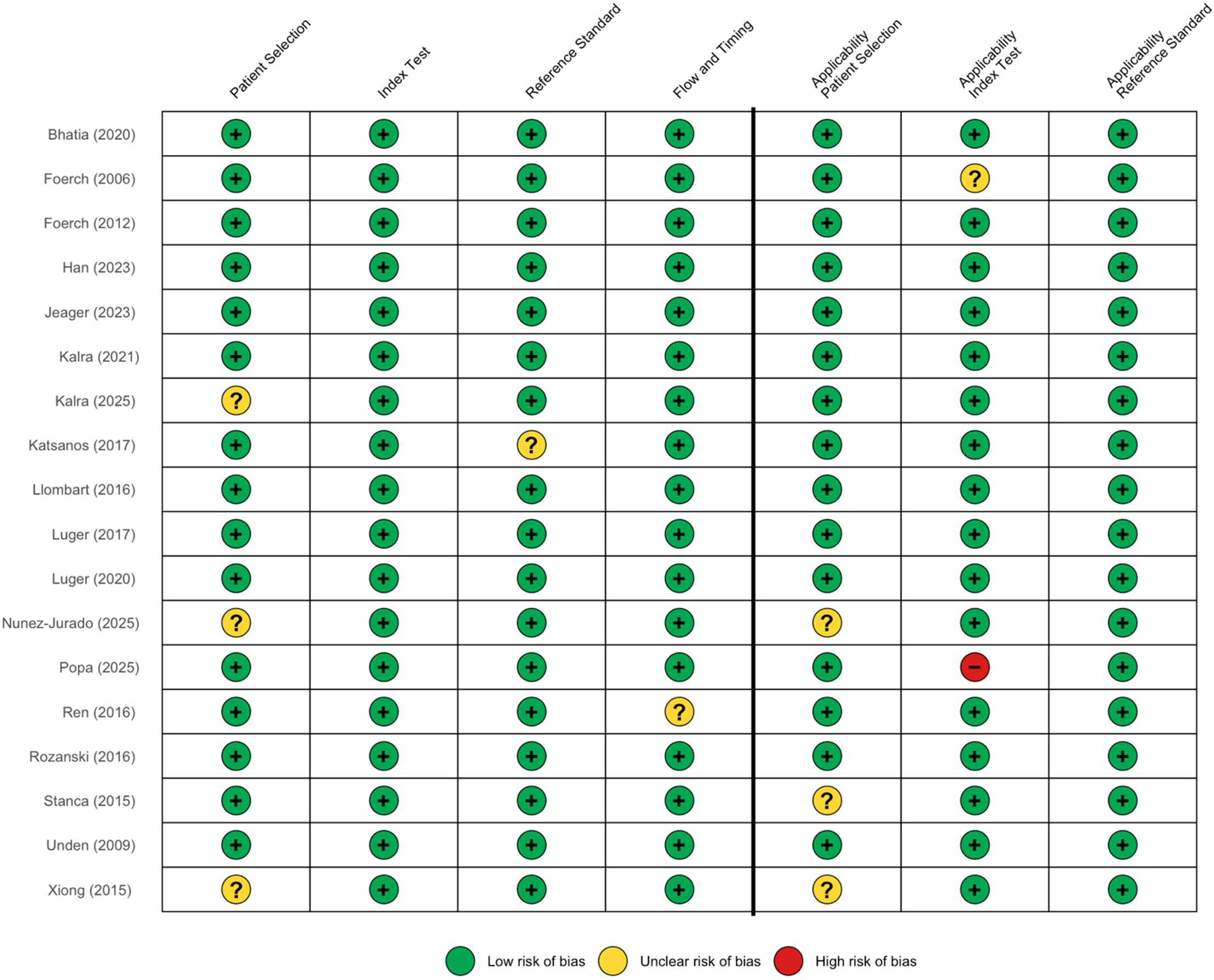
Risk of bias of studies included in the meta-analysis (n=18)

#### b. Study Design

The majority of studies included in the meta-analysis were prospective cohort studies (16, 88.9%), while 2 studies (11.1%) were retrospective cohort studies. Among the 17 studies reporting only associations with clinical and/or radiological outcomes, 12 were prospective cohort studies, 3 prospective cross-sectional, and 2 case-control in design.

A total of 45 out of 64 studies defined a time from symptom onset to hospital admission as part of study inclusion criteria. Among the 18 studies included in the meta-analysis, 2 selected patients who were admitted <4.5 hours, 9 studies <6 hours, 2 studies <12 hours, and 3 studies <24 hours while 2 studies did not have pre-defined time windows from onset to admission for participant inclusion.

#### c. Study Population

Study sample sizes ranged from 10 to 743 participants. Across studies, 10,600 participants were included, of which 1,403 were ICH, 6,555 were AIS, 67 were TIA, 572 stroke mimics and 1,365 healthy controls. The remaining participants had an unspecified stroke type, or served as non-stroke comparison participants in studies evaluating GFAP for stroke diagnosis among patients presenting with other conditions. Among AIS participants, at least 744 large vessel occlusion strokes (LVO) were identified, although LVO presence was not specified in all studies. Among all patients, at least 482 had a previous history of stroke.

Median study-specific NIHSS scores ranged from 2 to 15 across the 64 studies. More precisely, median NIHSS ranged from 3 to 16 in participants with AIS and 4.5 to 19.5 in those with ICH. Due to heterogeneity in reporting patient characteristics, a pooled estimate was not calculated for NIHSS.

### 3. Sampling Time, GFAP Measurement Methods, & Reference Standards

#### a. Sampling Time

Among the 64 studies, time from stroke onset to sampling was specified in 50 (78.1%) studies, and time from last known well (LKW) to sampling was reported in 8 (12.5%). All studies included in the meta-analysis specified time from onset to sampling. Of note, 2 studies reporting diagnostic accuracy data were excluded from the meta-analysis on account of: 1) one study reporting GFAP measurements at multiple specific time points rather than within predefined sampling windows, with no single time point that could be selected to represent the study (10) and 2) the other study assessed rates of change in GFAP levels.(14)

A total of 9 out of 64 studies (14.1%) reported inclusion of patients with an unknown time of stroke onset, whereas 3 (4.7%) stated their exclusion. The remaining studies (52 (81.3%)) did not report whether stroke of unknown onset were included or excluded.

Repeated GFAP sampling was performed in 17 (26.6%) studies, most commonly to evaluate associations of serial GFAP measurements with clinical outcomes.

#### b. GFAP Measurement Methods

GFAP measurement methods were heterogeneous across the 64 studies, with 39 (60.9%) studies using Enzyme-Linked Immunosorbent Assay (ELISA), 14 (21.9%) using Single Molecule Array (SIMOA) methods, 5 (7.8%) using electrochemiluminescence (ECL), 1 (1.6%) using Biochip Array Technology (BAT), 3 (4.7%) using chemiluminescent immunoassays (CLIA/CLEIA/CMIA) and 2 (3.1%) using a proteomics platform. Among the 18 studies included in the meta-analysis, ELISA was most frequently used (8, 44.4%), followed by ECL (4, 22.2%), and SIMOA (3,16.7%).

The 64 studies reported GFAP in ng/mL (30, 46.9%), pg/mL (29, 45.3%), ng/L (1, 1.6%), ug/L (1, 1.6%) or mcg/mL (1, 1.6%), while 2 (3.1%) studies reported relative expression values. For the meta-analysis, all GFAP values were converted to ng/mL for uniformity.

#### c. Reference Standard

A total of 59 (92.2%) studies specified the use of CT scan to confirm stroke diagnosis, of which 30 used both CT and MRI. One study performed MRI alone to confirm stroke diagnosis, while the imaging modality used was not specified in 4 studies.

### 4. Diagnostic Accuracy of Blood GFAP levels in Acute Stroke

#### a. Diagnostic Accuracy of GFAP to Differentiate ICH from AIS

A total of 12 studies evaluated the diagnostic performance of GFAP in differentiating ICH from AIS. Overall, the pooled HSROC-derived AUC was 0.86 (95% CI: 0.76-0.92), indicating good overall diagnostic accuracy of blood GFAP for differentiating ICH from AIS. (Figure 3) The pooled sensitivity of blood GFAP levels to differentiate ICH from AIS within 24 hours from stroke onset was 74.4% (95% CI: 63.2-84.2%) and the pooled specificity was 87.2% (95% CI: 77.0-94.7%). (Figure 4) Between-study heterogeneity was quantified using the Zhou–Dendukuri bivariate I² statistic and was estimated at 37.0%. Deeks’ funnel plot asymmetry test did not show significant asymmetry among the 12 studies (p = 0.195), suggesting no strong evidence of publication bias. (Figure 5)

**Figure 3:**
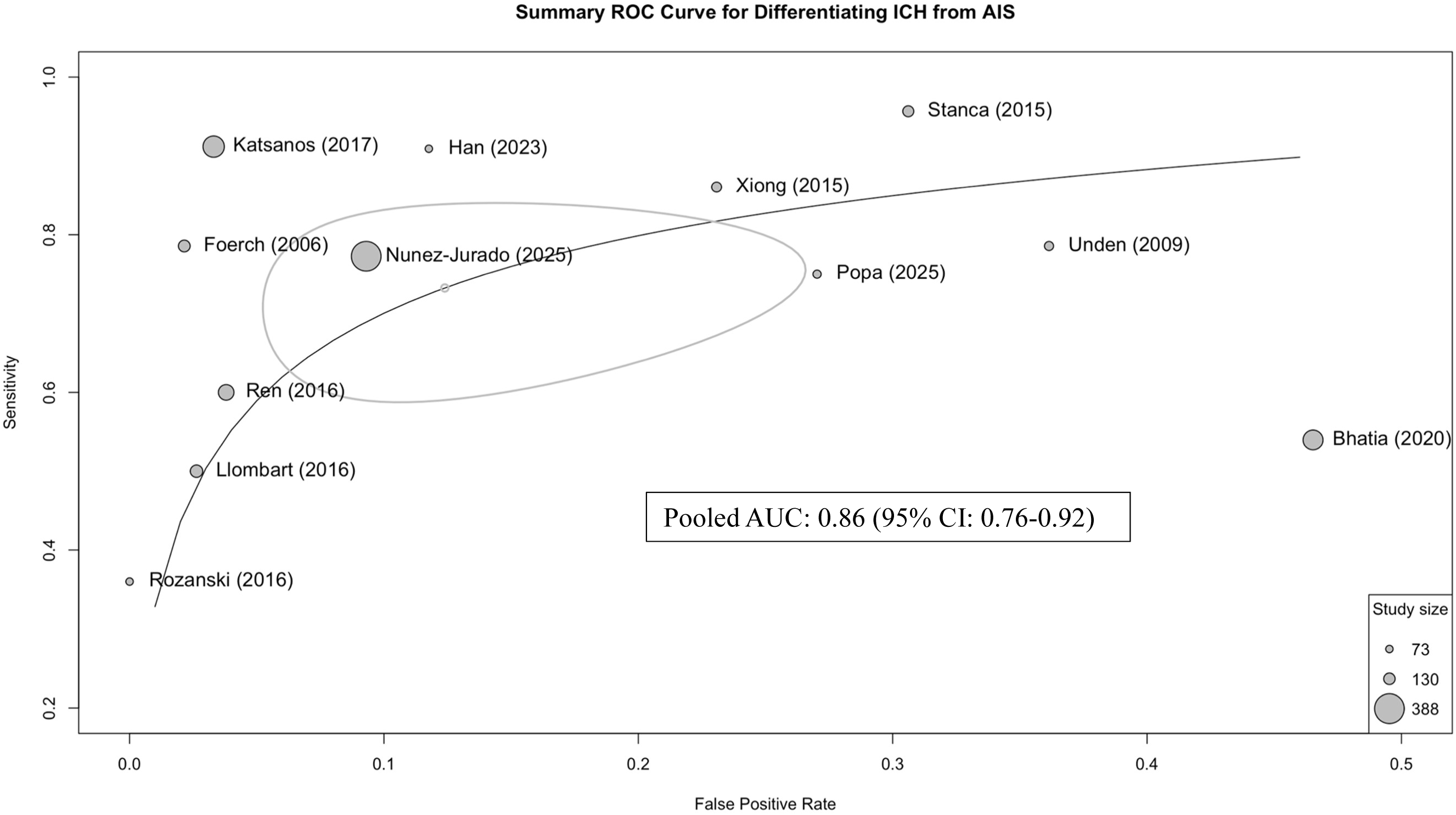
HSROC curve of studies assessing GFAP to differentiate ICH from AIS.

**Figure 4:**
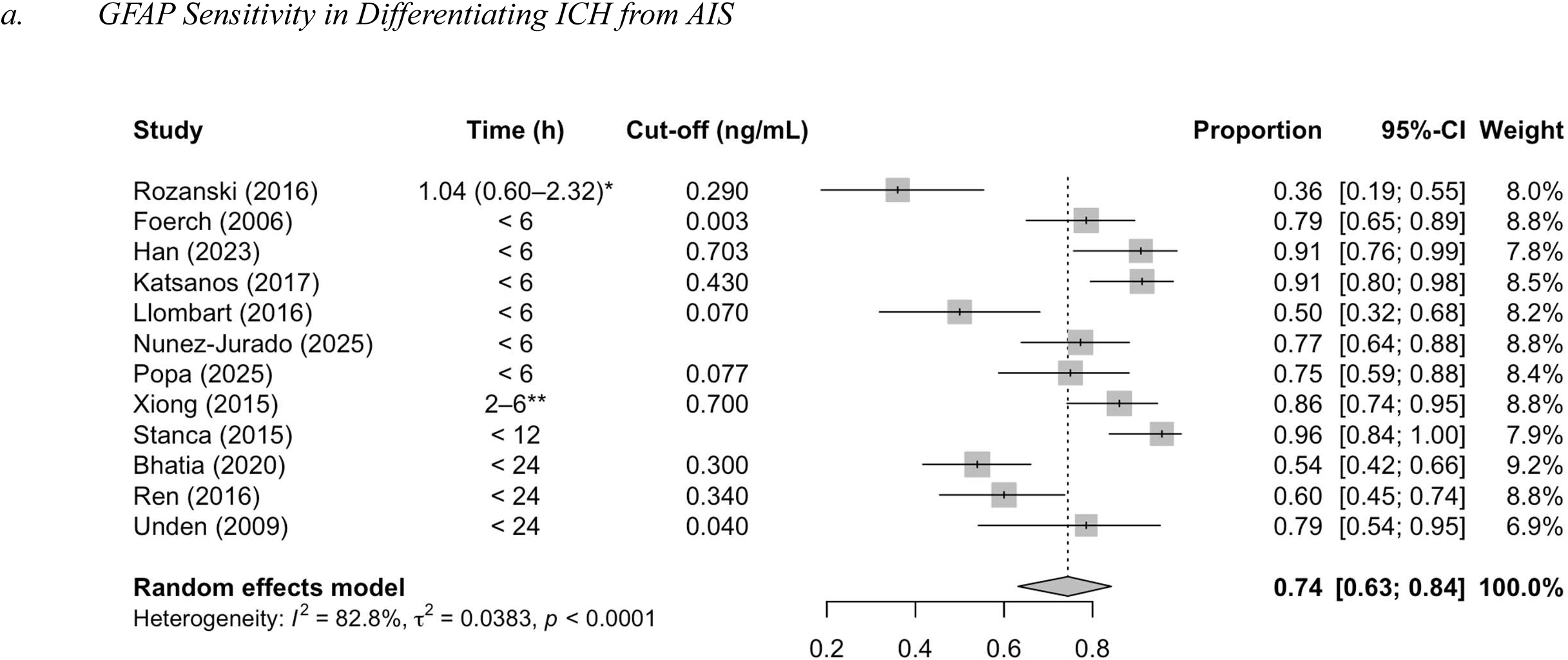
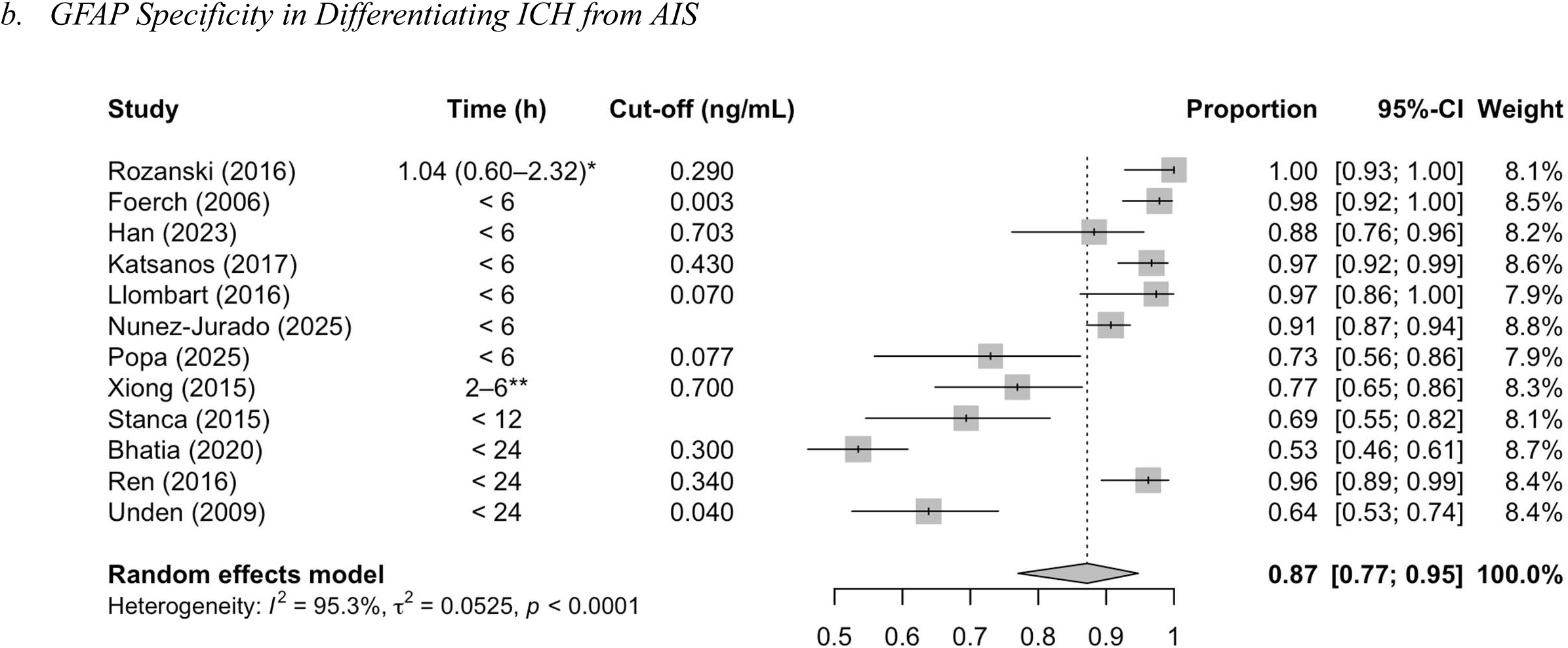
Forest plots for sensitivity (a) and specificity (b) of GFAP in differentiating ICH from AIS. Studies are presented in ascending order, from the shortest to longest time from symptom onset to sampling as a means of illustrating how sensitivity and specificity of GFAP vary as a function of time. Cut-offs were selected based on Receiver-Operating Characteristics (ROC) analysis from individual study data, or according to selected cut-offs from previous studies. * = Median (IQR), ** = Range

**Figure 5:**
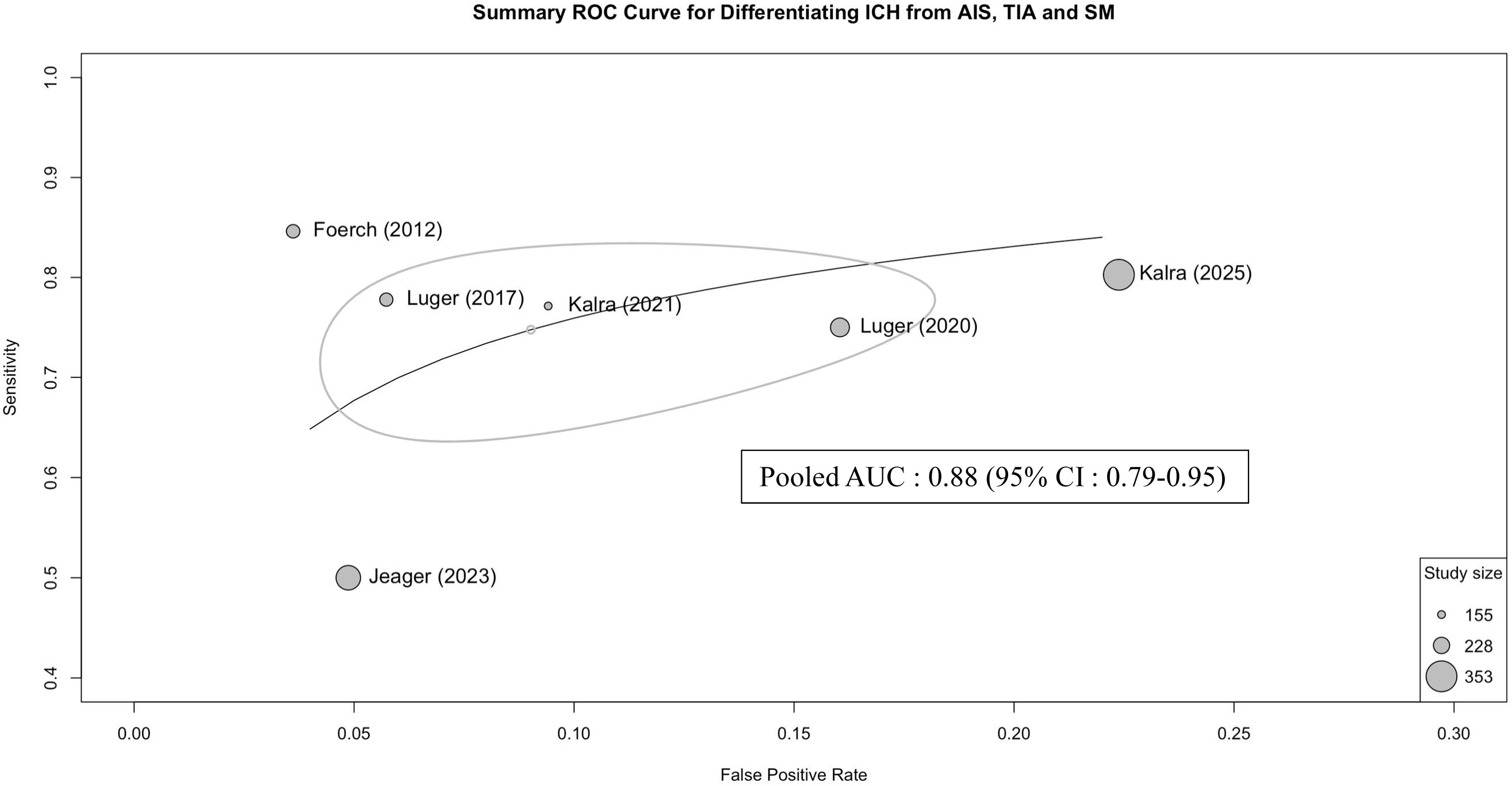
HSROC curve of studies assessing GFAP to differentiate ICH from undifferentiated suspected stroke (including AIS, TIA and stroke mimic)

Subgroup analyses were performed to evaluate GFAP diagnostic accuracy to differentiate ICH from AIS in studies measuring GFAP <6 hours from onset (n= 8 studies). The pooled sensitivity was 75.0% (95% CI: 61.1–86.7%), and the pooled specificity was 92.7% (95% CI: 84.9-97.8), with a corresponding AUC of 0.91 (95% CI: 0.81-0.96).

#### b. Diagnostic Accuracy of GFAP to Differentiate ICH Among Suspected Undifferentiated Suspected Stroke

Six studies evaluating GFAP’s diagnostic accuracy to differentiate ICH from all subtypes of undifferentiated suspected stroke (including AIS, TIA and stroke mimic) were identified. Blood samples were obtained <12 hours from onset in all studies (with none beyond 12-24h), including 2 in which sampling occurred within the first 4.5 hours.(28) Among these studies, the pooled HSROC-derived AUC was 0.88 (95% CI: 0.79-0.95). GFAP showed a pooled sensitivity of 75.4% (95% CI: 67.2-82.7%) and a pooled specificity of 90.5% (95% CI:84.1-95.4%). Between-study heterogeneity was quantified using the Zhou–Dendukuri bivariate I² statistic and was estimated at 41.2%. Overall, these findings indicate similarly good overall diagnostic accuracy of blood GFAP for differentiating ICH from all subtypes of suspected acute stroke (AIS, TIA, and stroke mimic), compared with its diagnostic performance for differentiating ICH from AIS alone. (Figures 5, 6)

**Figure 6:**
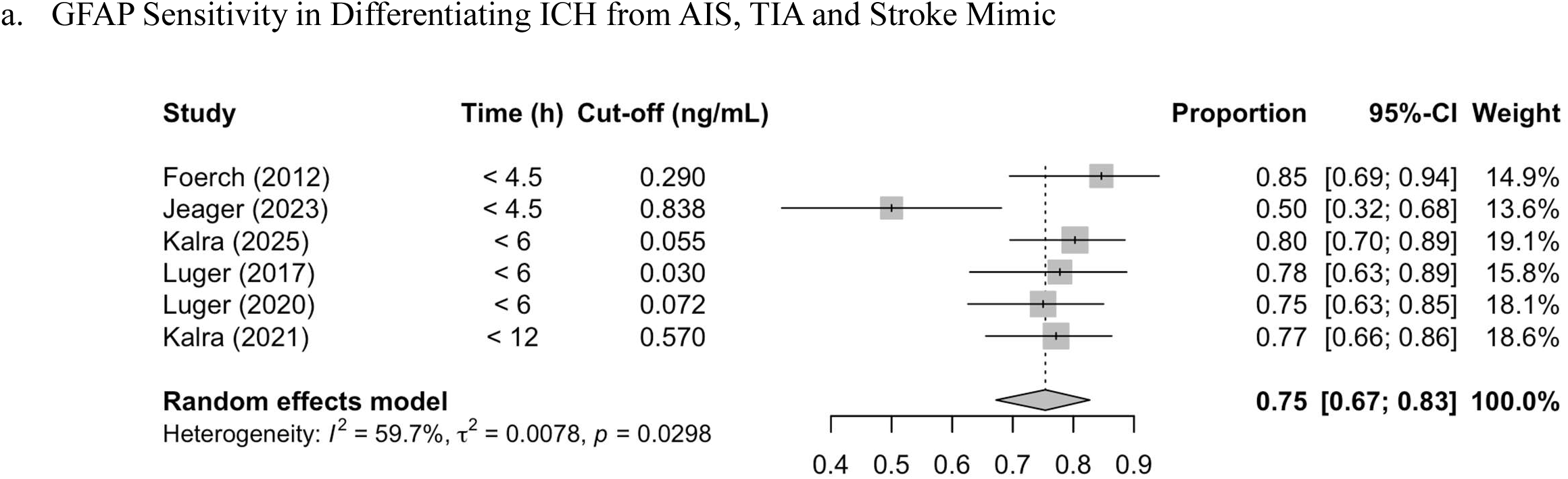
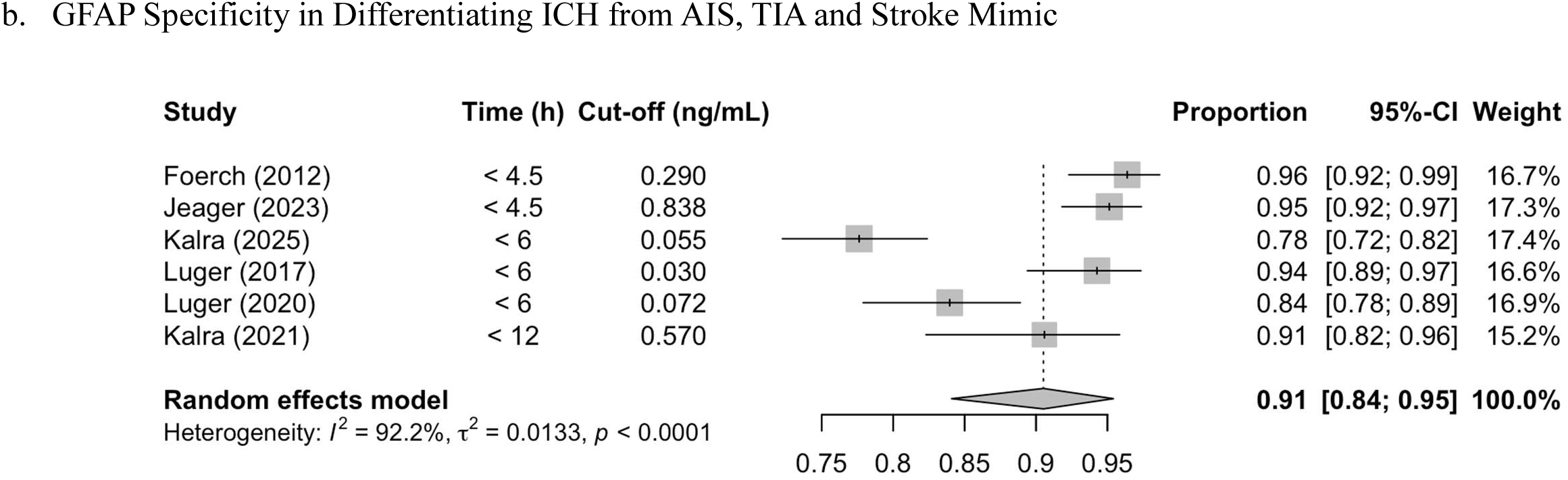
Forest plots for sensitivity (a) and specificity (b) of GFAP in differentiating ICH from undifferentiated suspected stroke including AIS, TIA and stroke mimic for each study. Studies are presented in ascending order, from the shortest to longest time from symptom onset to sampling as a means of illustrating how sensitivity and specificity of GFAP varies as a function of time. Cut-offs were selected based on Receiver-Operating Characteristics (ROC) analysis from individual study data, or according to selected cut-offs from previous studies.

#### c. Diagnostic Accuracy of GFAP in the Ultra-Early Phase of Stroke

Subgroup analyses were performed to evaluate GFAP diagnostic accuracy within 2h from onset (n=5 studies). Given the small number of studies, those assessing ICH from AIS were pooled together with studies assessing ICH from suspected acute stroke for analyses. GFAP levels in this ultra-early time window had a pooled sensitivity of 76.7% (95%CI: 66.8-85.3%), a pooled specificity of 96.4% (95% CI: 93.9-98.2%), with a corresponding AUC of 0.96 (95% CI: 0.80-0.97%).

### 5. Clinical and Radiological outcomes

#### a. GFAP and Stroke Severity (NIHSS)

A total of 18 studies assessed the association between stroke severity (as assessed by admission NIHSS score) and GFAP concentrations in acute stroke. (Supplementary Table 2) Of the 18 studies, 17 included AIS participants, 7 included ICH, and 6 included both stroke types. A total of 11 studies reported a significant association between GFAP values and NIHSS in AIS, of which 8 studies identified a positive correlation between GFAP and admission NIHSS. Five studies reported no significant association between GFAP and NIHSS in AIS. In ICH, 5 reported a significant association between admission NIHSS and GFAP, of which 4 demonstrated a positive correlation, and 2 found no association.

#### b. GFAP and the Extent of Ischemic Damage

Three studies (4.3%) assessed the relationship between the extent of ischemic brain tissue injury (as per the ASPECTS score) and GFAP levels at admission in AIS (Supplementary Table 3). Among these studies, higher GFAP levels were found to be significantly associated with a lower ASPECTS score (suggesting more extensive ischemic injury) on baseline neuroimaging in 2 studies.(15) (13) (16)

#### c. GFAP and ICH Volume

Twelve studies (17.4%) assessed the relationship between GFAP concentrations and ICH volume. (Supplementary Table 4) Of these, 10 studies reported a positive correlation with ICH volume, with correlations coefficients ranging from moderate to strong (0.45-0.84, all p<0.05). Importantly, 7 of the 12 studies reported that small ICH volumes (generally <10 ml) were associated with low GFAP levels or values below the assay’s limit of detection, while larger hematoma volumes were consistently associated with higher GFAP values.(10–12,21,26,29,30) Two studies also reported ICH location as a potential modifier of this relationship, with lobar ICH showing higher GFAP values than brainstem or deep locations. (31)

## DISCUSSION

Interest in blood-based biomarkers for acute stroke triage has grown substantially, driven by the need for rapid diagnostic tools suited to prehospital and resource-limited settings. Blood GFAP has emerged as a promising candidate. This systematic review and meta-analysis provide an updated summary of the literature regarding the diagnostic performance of GFAP in acute stroke, particularly in differentiating ICH from AIS, TIA and stroke mimics.

Across the identified studies, potential sources of bias included inadequate reporting of participant enrollment, and inappropriate exclusions that may have overrepresented patients with more severe strokes. Study populations were also heterogeneous, and time from stroke onset was reported inconsistently across studies.

Nevertheless, across 18 studies selected for the meta-analysis, risk of bias was low overall, and GFAP demonstrated value as an early diagnostic biomarker in acute stroke. The pooled AUC of GFAP in differentiating ICH from AIS within 24 hours from stroke onset was 0.86, with a pooled sensitivity of 74.4% and pooled specificity of 87.2%. Diagnostic performance of GFAP to differentiate ICH from other subtypes (AIS, TIA and stroke mimics) was similar, with an AUC of 0.88, a pooled sensitivity of 75.4% and a pooled specificity of 90.5%. These findings are consistent with those from a previous meta-analysis, in which pooled sensitivity and specificity were 75.6% and 94.5%, respectively. (32) Across both comparisons, GFAP’s higher specificity relative to sensitivity supports its use primarily as a rule-in test for ICH detection in the acute setting of suspected stroke. Indeed, rapid ICH detection could support earlier initiation of blood pressure reduction in the ambulance, potentially improving ICH outcomes as suggested in the recent INTERACT 4 study. (3)

Current literature suggests that the timing of GFAP sampling from stroke onset may influence the biomarker’s diagnostic performance in acute stroke. In our updated meta-analysis, specificity remained high among the studies assessing GFAP diagnostic accuracy <6 hours from stroke onset, consistent with prior studies showing that GFAP performs well within this time window. (9) Similar patterns of high specificity have also been reported in subgroups <3 hours from stroke onset(32).

In contrast, few studies specifically reported GFAP performance <2 hours from onset - a time window most relevant to potential out-of-hospital application. (30) Our subgroup analyses in this time window revealed that GFAP retains high specificity and high point-estimate AUC, a finding which may be clinically relevant, where decisions regarding early intervention in ICH, such as early BP treatment are important, and the risk of misclassifying ICH as AIS are most severe. The high specificity observed supports GFAP’s potential for reliable early identification of ICH. Nevertheless, it is important to highlight that this analysis was based on few studies (n=5), necessitating the pooling of studies assessing ICH vs. AIS with those assessing ICH against a broader undifferentiated suspected stroke population. Given the small number of contributing studies, the precision of this pooled estimate should be interpreted with caution, and between-study heterogeneity could not be reliably assessed in this subgroup.

In our subgroup analysis, GFAP also demonstrated a pooled sensitivity of 76.7% within the first 2 hours from onset, comparable to the pooled sensitivity estimated across all studies. Nevertheless, previous literature suggests that sensitivity may be comparatively limited within the first two hours from onset, which may compromise its clinical utility as a standalone rule-out test. In the recent DETECT study, sensitivity within 2 hours from onset ranged from 36.8% to 65.0%, with sensitivity varying with age, while specificity remained high (98.1–98.3%).(21) Of note, these data could not be included in our subgroup analyses within the 2h time strata given that data in this time window was not pooled due to age-stratified reporting without a combined estimate. Another study, which measured GFAP at multiple specific timepoints, similarly reported high specificity (100%) but low sensitivity (45%) for differentiating ICH from AIS at 2 hours from onset.(10)

Several factors may contribute to this reduced sensitivity. From a pathophysiological perspective, early astroglial injury secondary to hematoma formation may be insufficient to generate substantial circulating GFAP levels, at earlier timepoints with peak GFAP concentrations occurring at approximately 2 hours after ICH onset according to a modeled estimate.(23) This is supported by results from our narrative review of studies. Among the 12 studies that reported GFAP levels in relation to hematoma volume, 11 showed a moderate-strong correlation between GFAP levels and ICH volumes, while 7 studies specifically noted that smaller hematomas (typically <10 mL) were associated with GFAP values often below limits of detection, highlighting the possibility of false-negative results in this subset particularly in the first 2 hours from onset.

Larger, prospective studies specifically designed to evaluate GFAP kinetics within this critical ultra-early window are needed to confirm these findings and to determine optimal diagnostic thresholds for prehospital use. Of note, one study demonstrated that measuring serial prehospital sampling of GFAP with measurements of GFAP release rates may overcome the limited sensitivity in the ultra-early phase of stroke, further underscoring the time-dependent nature of GFAP as a diagnostic biomarker. (14) A prior meta-analysis also suggested the use of a combination of biomarkers spanning different biological pathways could improve diagnostic accuracy. (9) The emergence of point-of-care (POC) assays allowing rapid GFAP measurements, along with future studies in the prehospital setting, may improve characterization of GFAP diagnostic accuracy in earlier time windows.

Few studies (n=3, 4.3%) evaluated the relationship between admission GFAP levels and ischemic injury extent, as measured by the ASPECTS score. (15) (13) (16) Two of the three reported that higher GFAP levels were significantly associated with lower ASPECTS scores, consistent with greater astrocytic injury and GFAP release in more extensive ischemic damage. Nonetheless, given the small number of studies, these findings are hypothesis-generating rather than conclusive, and larger studies are needed to confirm whether GFAP reliably informs the extent of ischemic burden in AIS, particularly in patients with delayed or unknown onset.

### Limitations

Several limitations of our study warrant consideration. Firstly, in the studies identified in the literature, serial GFAP sampling was infrequently performed. Although GFAP levels are known to vary significantly over time in acute stroke (6), few studies provided GFAP diagnostic performance data at multiple timepoints. Thus, the ability to model the biomarker’s dynamic release pattern according to stroke subtype remains currently limited. Time intervals to sampling were also measured from various clinical reference points other than symptom onset, such as last known well or time of admission, and were expressed as either mean or median values, affecting comparability across studies.

Secondly, GFAP assay technologies varied considerably, with 60.9% studies using Enzyme-Linked Immunosorbent Assay (ELISA) and 21.9% using Single Molecule Array (SIMOA). Of note, SIMOA is known to be more sensitive than ELISA for GFAP detection, which may have impacted the overall diagnostic accuracy of the meta-analysis.(28,33) Furthermore, diagnostic cut-offs and their selection, which was based either on previous literature or derived from individual study data to maximize sensitivity and specificity, were inconsistent across studies. Identification of a standardized cut-off value to differentiate stroke subtype is recommended in future studies in order to guide clinical implementation for GFAP use. (8) Additionally, between-study heterogeneity was moderate (I² = 37.0%), which may partly reflect the variability in assay platforms, diagnostic cut-offs, and timing of sample collection discussed above. While Deeks’ funnel plot asymmetry test did not show significant evidence of publication bias (p = 0.195), the limited number of included studies (n=12) likely reduced the power of this test, and the possibility of unpublished negative studies cannot be fully excluded.

Thirdly, study populations were heterogeneous with respect to included stroke subtypes. Stroke mimics notably comprised only 5.4% of the pooled population, with an expected rate of 24.8% among participants presenting with suspected ischemic stroke according to a comprehensive review.(34) This underrepresentation may affect the external generalizability of results to a broader population presenting with undifferentiated suspected stroke.

Fourth, few studies specified whether stroke of unknown onset were included or excluded from analyses, thereby reducing external validity in the frequent clinical scenario where time of stroke onset is unknown, particularly among patients presenting with wake-up stroke or delayed (up to 24h) presentations. Biomarker-guided selection could be particularly relevant to determine treatment eligibility in these subgroups of patients.

Furthermore, associations between GFAP and clinical or radiological outcomes were derived from studies with heterogeneous designs and statistical approaches. Coupled with the exploratory nature of these analyses, these findings were narratively summarized and should be interpreted as hypothesis-generating.

Lastly, while not an *a priori* exclusion criterion, this review was limited to studies published in English or to studies in which translation of the text was deemed reliable, thus potentially omitting relevant data.

## CONCLUSION

This systematic review and meta-analysis provides a comprehensive and updated synthesis of GFAP’s diagnostic performance in differentiating ICH from AIS, TIA, and stroke mimics. Across 18 studies included in the meta-analysis, GFAP showed good diagnostic performance for early differentiation of ICH from AIS (AUC 0.86, pooled sensitivity 74.4%, specificity 87.2%) and from undifferentiated suspected stroke (AUC 0.88, sensitivity 75.4%, specificity 90.5%). AUC and specificity increased in earlier sampling windows, reaching 0.96 and 96.4% within 2 hours of onset, respectively. Sensitivity remained comparatively limited across all time strata (74.4–76.7%), likely reflecting lower GFAP values in patients with small ICH volumes, where limited parenchymal injury may fall below the threshold for reliable biomarker detection. These findings indicate that GFAP has strong rule-in, rather than rule-out, value for identifying ICH in the acute setting, as a negative result cannot reliably exclude ICH of small volume that may nonetheless be clinically significant, particularly in prehospital or resource-limited settings where immediate neuroimaging is unavailable. Furthermore, knowledge gaps persist regarding the potential of GFAP to assess ischemic injury extent in AIS, particularly among those with delayed or unknown onset.

## Data Availability

This systematic review was registered with PROSPERO (#CRD42024608605)prior to data extraction. Data extracted for this systematic review/meta-analysis are derived from previously published studies which are cited within the manuscript. The dataset used for the meta-analysis is available from the corresponding author upon reasonable request.

